# Sleep quality mediates the relationship between risk of obstructive sleep apnea and acute stress in young adults

**DOI:** 10.1101/2021.03.11.21253368

**Authors:** Kunal Aggarwal, Nasreen Akhtar, Hrudananda Mallick

## Abstract

**Purpose:** Intermittent hypoxia and transient arousals in obstructive sleep apnea (OSA) can lead to poor sleep quality and acute stress. Rising levels of obesity and increased incidence of OSA in young adults predisposes them to acute stress. We propose a mediation model to assess if risk of OSA is associated with acute stress and if the relationship between risk for OSA and acute stress is mediated by sleep quality.

**Methods:** 493 healthy individuals (F=237, M=256) from 18-25 years of age (mean age=20.3±1.53 years) were screened for OSA, sleep quality and acute stress using STOP-BANG questionnaire, Pittsburg Sleep Quality Index and American Psychiatry Association’s National Stressful Events Survey Acute Stress Disorder Short Scale (NSESS-S) respectively. Binary and logistic regression were used establish the relationships between the variables. Sobel test for mediation analysis was conducted.

**Results:** 73 participants (17.3%) were found at an intermediate and high risk of OSA by STOP BANG questionnaire. 79 (16%) participants reported level of stress as ‘None’. Mild, moderate and severe stress was present in 248 (50.3%), 109 (22.1%), 51 (10.3%) and 16 (3.2%) participants respectively. The odds of having severe and extreme stress among those at risk of sleep apnea is 2.18 times higher than that among those not at risk of sleep apnea (OR: 2.18, 95% Confidence Interval: 1.37-3.51). Sobel test established that the relationship between OSA and acute stress is mediated by sleep quality.

**Conclusion:** Sleep quality mediates the relationship between risk for sleep apnea and acute stress. This highlights the importance of screening for OSA in young adults, particularly young men with high BMI, presenting with high stress levels.

## Introduction

Obstructive sleep apnea (OSA) is characterized by repeated upper airway obstruction causing oxygen desaturation and frequent arousals from sleep[1]. The prevalence of OSA is increasing worldwide. A study from India reported a prevalence of 13.74% of OSA in general population[2]. The frequent desaturations, electroencephographic microarousals and sleep fragmentation in OSA lead to cardiovascular and neuropsychiatric sequalae [3]. OSA has an effect on intellectual abilities related mainly to verbal ability, attention and learning skills including planning and object categorisation [3]. These represent an organic damage to the central nervous system. The awareness of these intellectual impairments, sleep fragmentation and reduced daytime alertness makes the patients feel restless, anxious or depressed [4,5]. This can manifest emotionally as stress. Frequent arousals also cause an increase in catecholamines and cortisol levels, exacerbating the stress. Emotional stress has been found to be higher in patients of OSA and reduces after treatment [6]. OSA has been linked to quality of life [7], ^7^ depression, ^8^ and anxiety, ^9^ but the association with acute stress has been reported by very few studies. ^6,10^ Most of the studies on OSA have collected data from moderate to severe sleep apnea in older patients and in clinical settings. Occurrence of OSA in general population, particularly among young adults, has been neglected. Young adults are a vulnerable population for higher emotional stress. The current study was conducted in young adults in general population with the objectives to investigate the association of between risk of OSA and acute stress levels and to assess if the association between risk of OSA and acute stress level is mediated by sleep quality.

## Material and methods

### Sample and data Collection

This data is obtained from a study conducted in undergraduate college students from various courses in New Delhi from July to September 2019. ^11^ Ethical clearance was obtained from Institutional Ethical Committee of All India Institute of Medical Sciences, New Delhi (No IECPG-343/29.05.2019 dated 03.07.2019). The primary objective of the study was to investigate the association between acute stress and risk for obstructive sleep apnea in young adults.

This was a cross-sectional and observational study. There was no control group or blinding of assessors. Participants were approached by advertising in college social media groups and announcements in classrooms. The complete information regarding the study and consent form were provided online. All participants gave informed consent for the study and filled the online questionnaire. The questionnaire collected information about the student’s age, gender, height, weight, acute stress, risk for sleep apnea and sleep quality. 503 participants between the ages of 18 and 25 years consented for the study. Those who were previously diagnosed for or on treatment for sleep disorders, psychiatric disorders or any other chronic disease were excluded.

### Measures

Risk of sleep apnea was assessed by using STOP-BANG questionnaire (SBQ). ^12^ Even though Berlin questionnaire is the one most used in assessments in the clinic, SBQ is the more accurate tool for assessing risk of OSA in the general population. ^13^ There were 8-items in the questionnaire. Each positive item was scored as one. The risk was marked low, intermediate and high for a score of 0-2, 3-4 and 5-8 respectively. Intermediate and high risk categories were clubbed into one category for the purpose of analysis in this study as the number of participants in high risk category were very few and would skew the analysis.

Sleep quality was assessed by using Pittsburg Sleep Quality Index. ^14^ It is a 9-item questionnaire widely used for assessing sleep quality. A score of less than or equal to five was considered good quality sleep, and a score of more than five was considered as poor-quality sleep. Sleep time was distinguished from other activities after going to bed like listening to music or reading books. Acute stress was assessed by American Psychiatry Association Severity of Acute Stress Symptoms—Adult (National Stressful Events Survey Acute Stress Disorder Short Scale [NSESSS]) questionnaire. ^15^ It was a 7-item questionnaire. Each item on the measure was rated by the subject on a 5-point scale (0=Not at all; 1=A little bit; 2=Moderately; 3=quite a bit, and 4=Extremely). The total raw score ranged from 0 to 28 and a higher score indicated a higher degree of acute stress. This score was divided by 7 (the number of items) to provide a total average score which was allocated a 5-point scale. It was used to assess severity of acute stress as none (0), mild (1), moderate (2), severe (3) and extreme (4). ^15^

### Analysis

Acute stress, sleep quality and risk for obstructive sleep apnea were correlated to each other using Pearson’s bivariate correlation analysis. The Statistical Package for the Social Sciences (SPSS^®^) software version 25.0 (IBM, Armonk NY, USA) for Windows^®^ was used for the correlation analyses. To investigate if risk for sleep apnea is associated with acute stress and if sleep quality mediates the relationship between sleep apnea and acute stress, we created a mediation model (Figure 1). A simple mediation analysis was performed based on the method described by Baron and Kenny. ^16^ The dependent (outcome) variable for analysis was acute stress. The independent variable was risk for obstructive sleep apnea. Mediator variable was sleep quality. In our model, we assume there are two causal paths to the dependent variable (acute stress), the direct impact of risk of OSA (path c) and the impact of sleep quality as the mediator (path a and b). The mediated path from independent variable (Risk of OSA) to the dependant variable (acute stress) is path c’. Binary and ordinal logistic regression were performed using R software (R Core Team 2014 v3.4.4). Mediation analysis was performed using Sobel test. ^17^ Sobel test is an accepted test of indirect effect and mediation. ^18^

**Figure 1.**
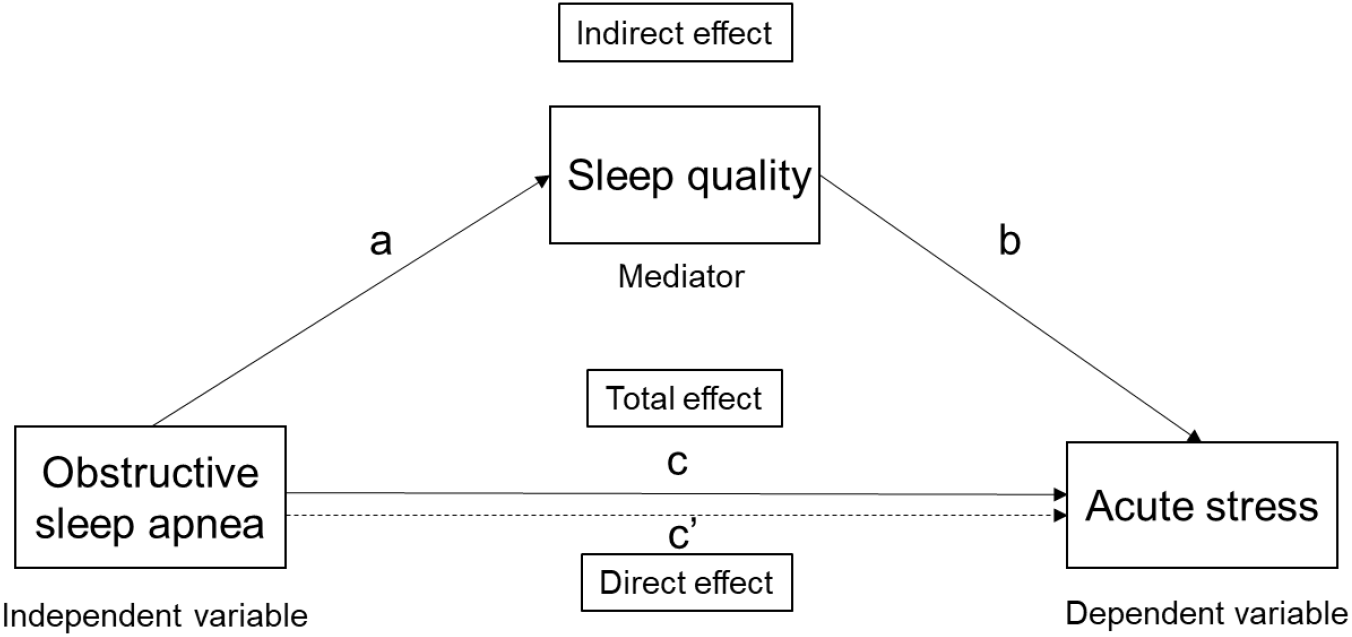
Mediation model

**Figure 2.**
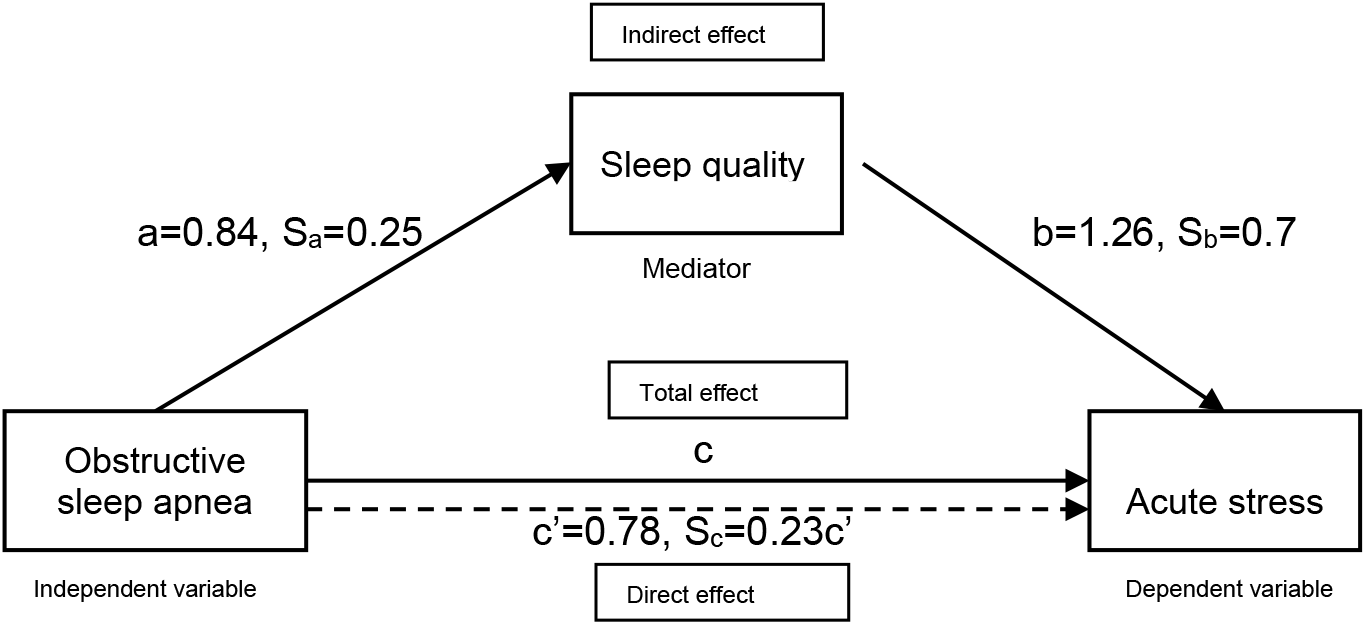
Mediation model with effect size. Legend: a, Regression coefficient of pathway ‘a’; b, Regression coefficient of pathway ‘b’; c, Regression coefficient of pathway ‘c’; S_a_, Standard error of the coefficient ‘a’, S_b_, Standard error of the coefficient ‘b’, S_c_, Standard error of the coefficient ‘c’

## Results

503 volunteers filled up the questionnaire. After applying inclusion and exclusion criteria, 493 were included for data analysis. Female: male ratio was 0.91 (Females= 237, Males = 256). Mean age of participants was 20.3±1.53 years, ranging from 18 to 25 years. The basic descriptives of the study participants are depicted in Table 1. 73 participants (17.3%) were found at an intermediate and high risk of OSA. 79 (16%) participants reported level of stress as ‘None’. The prevalence of mild, moderate and severe stress was 248 (50.3%), 109 (22.1%), 51 (10.3%) and 16 (3.2%) respectively. Mild stress was found higher in males (males-27.2%, females-23.1%), whereas moderate, severe and extreme stress was higher in females (Table 1).

**Table 1.** Basic descriptives of the study participants

| S.No | Characteristic | Total<br>(n=493) | Male<br>(n=256) | Female<br>(n=237) |
| --- | --- | --- | --- | --- |
| 1 | Age (years) Mean ± SD | 20.3±1.53 | 20.04±1.46 | 20.59±1.57 |
| 2 | BMI (Kg/m<sup>2</sup>) Mean ± SD | 22.7±3.77 | 22.97±3.49 | 20.44±3.91 |
| 3 Acute stress |  |  |  |  |
|  | None (0) | 69 (14%) | 43 (8.7%) | 26 (5.3%) |
|  | Mild (1) | 248(50.3%) | 134(27.2%) | 114(23.1%) |
|  | Moderate(2) | 109(22.1%) | 49 (9.9%) | 60(12.2%) |
|  | Severe (3) | 51(10.3%) | 25(5.1%) | 26(5.3%) |
|  | Extreme (4) | 16 (3.2%) | 5 (1.0%) | 11(2.2%) |
| 5 Risk of Obstructive Sleep Apnea |  |  |  |  |
|  | Low risk | 420 (85.2%) | 192 (75%) | 228 (96.2%) |
|  | Intermediate + High risk | 73 (14.8%) | 64 (25%) | 9 (3.7%) |
| 6 Sleep quality (PSQI) |  |  |  |  |
|  | Good (Score 1-5) | 315 (63.9%) | 173 (35.1%) | 142 (28.8%) |
|  | Poor (Score 5- 21) | 178 (36.1%) | 83 (16.8%) | 95 (19.3%) |

**Notes:**
**Abbreviations:** SD, Standard Deviation, PSQI, Pittsburg Sleep Quality Index

### Correlation analysis

Acute stress was significantly correlated with sleep quality (r=0.328, p<0.001), and risk for OSA (r=0.160, p<0.001) (Table 2). Sleep quality also correlates with risk of OSA (r=0.150, p<0.001).

**Table 2:** Correlation between acute stress, Sleep Quality and Risk of OSA

|  | Acute stress | Sleep quality | Risk of OSA |
| --- | --- | --- | --- |
| Acute stress | 1 | .328** | .160** |
| PSQI |  | 1 | .150** |
| SB |  |  | 1 |

Abbreviations: OSA, Obstructive Sleep Apnea

### Logistic Regression and mediation analysis

The assumptions of regression were checked and found satisfactory. First, using steps described by Baron and Kenny, ^16^ using acute stress as the dependant variable and risk for obstructive apnea as the independent variable, ordinal logistic regression was applied and risk for obstructive apnea was shown to be a significant predictor (p<0.001) of acute stress for all levels of stress (none, mild, moderate, severe and extreme) (c pathway). The odds of having severe and extreme stress among those among those at risk of sleep apnea is 2.18 times higher than that among those not at risk of sleep apnea (OR: 2.18, 95% Confidence Interval: 1.37-3.51). Second, risk for sleep apnea was used to predict the mediator variable of sleep quality (a pathway) using binary logistic regression. The odds ratio of poor sleep quality was 2.3 times in a person at high risk of OSA compared to a participant at low risk of OSA (OR: 2.3, 95% Confidence interval: 1.40-3.83). Third, the relationship between the mediator sleep quality and acute stress was established using ordinal logistic regression (b pathway). On change in sleep quality from good to poor, the odds of mild stress vs moderate or severe combined are 3.5 times greater (OR: 3.55, 95% Confidence interval: 2.48-5.11). Last, the mediated relationship between SBQ and acute stress was examined for a reduction in prediction when the mediator was added to the model (c’ pathway). Based on a previous study, ^19^ the three regression equations were used to test the mediation effects of SQ on relation between OSA and acute stress. Utilizing the coefficients of pathways ‘a’ and ‘b’, and Standard Errors (S_a_, S_b_) of these coefficients, mediation was found (Test statistic: 0.7255; p-value 0.049, using Sobel test), [8]^7^ showing that the relationship between SB and acute stress was no longer significant after controlling for sleep quality. The Sobel test was used to determine that the ‘ab’ effect was significantly greater than zero. This provides evidence that the relationship between OSA and acute stress is mediated by sleep quality.

## Discussion

Recurring upper-airway obstruction, intermittent hypoxemia and transient arousals leading to sleep fragmentation and poor sleep quality, are the hall marks of OSA. The risk factors for OSA are male sex, obesity, and advancing age, among others. ^6^

Rate of weight gain is greatest in young adults, averaging 1 to 2 pounds per year. ^20^ Up to 19% of teenagers and young adults are reportedly obese in India. ^21^ Rising levels of obesity are increasing the prevalence of OSA in young adults. The role of OSA in development of stress in young adults has not been studies much. When presenting with acute stress, young adults are seldom screened for OSA. In this study, our aim is to highlight the importance of screening for OSA in young adults, particularly young men with high BMI, presenting with high stress levels. SBQ has been recommended as the screening tool for OSA in community settings and where PSG is not available. ^13^ It is more accurate than Berlin and STOP questionnaire for screening of OSA in general population. ^13,22^ SBQ has a reported sensitivity of is 88,90 and 93%, specificity of 42,36 and 35%, and a diagnostic odds ratio of 5.13, 5.05 and 6.51 for mild, moderate and severe OSA, respectively. ^13^ We found a prevalence of intermediate and high risk of sleep apnea to be 14.8% in our study. It is similar to the prevalence rate of 13.7% reported by a previous study conducted in Delhi. ^2^ We have found a significant association of risk of OSA with acute stress. We presented the result in World Sleep Congress 2019. ^23^ The risk of being under severe acute stress was 2.18 times higher in those at risk of OSA than those without the risk of OSA. Other studies have evaluated the association of OSA with emotional stress. Das Santos et al have reported a higher stress level in moderate and severe apnea patients, which reduced significantly after receiving medical or surgical treatment.^6^ Frequent arousals during the night in OSA alter the activity of hypothalamic-pituitary-adrenal axis. Repeated and reversible airway obstruction lead to intermittent hypoxia, sleep fragmentation and decreased sleep duration, causing cortisol and catecholamine release. ^24–27^ Cortisol release is associated with higher levels of stress. ^28^ This may be contributing to acute stress development in patients of OSA.

Second, we assessed if risk for OSA predicts the quality of sleep. As expected, we found higher odds of poor-quality sleep in those at high risk of OSA as compared to those at low risk of OSA. Poor quality of sleep is one the major symptoms of OSA and causes excessive daytime sleepiness and fatigue. ^1^ Quality of sleep is closely related to number and extent of arousals. Frequent arousals are a hallmark of OSA. ^1,9^

Third, we assessed if sleep quality can predict acute stress. Using ordinal logistic regression, we found that poor sleep quality leads to 3.5 times higher chance of having moderate/ severe acute stress. Some other studies have found that increased perceived stress and sleep quality are associated with each other. ^29,30^ Doane and Thurston found that lower sleep efficiency predicted higher next day stress in adolescents. ^31^ A few studies have examined the directionality of the relation between stress and sleep and found it to be bidirectional. ^27,31^ Sleep quality affects the quality of replenishment of cognitive processes and energy level. ^32^ Experimentally produced sleep restriction or sleep loss produces performance decline the next day. ^32,33^ Our study is unable to comment on causality or directionality of effect due to a cross-sectional design. The importance of our study lies in relating the sleep quality in everyday settings of the participants to acute stress.

Mediation analysis proves our hypothesis that poor sleep quality mediates the relationship between OSA and acute emotional stress. Repeated inspiratory airway occlusion in OSA leads to multiple stressors. The stressors may be frequent episodes of hypoxemia of varying duration and severity, disrupted sleep due to frequent arousals, reduced total sleep time, and strenuous respiratory efforts resulting in the generation of severe negative intrathoracic pressures. ^35^ The neuro-hormonal, vascular and endocrine responses to OSA include excessive sympathetic activation which may carry over to the daytime awake state. ^36^ The powerful sympathetic excitatory effects may cause increased glucocorticoids and catecholamines and translate into acute stress. ^37^

One of the limitations of our study is the inability to establish the diagnosis of OSA with polysomnography. Further studies using polysomnography for diagnosis of OSA with objective measures of stress such as cortisol levels are required to confirm the findings. Secondly, the duration and onset of acute stress and OSA cannot be determined due to cross sectional design, and hence the temporal association of acute stress with OSA cannot be established.Based on the findings of our study, we recommend screening for OSA in young adults presenting with acute stress, particularly in overweight males. Further studies using polysomnography for confirmatory diagnosis of OSA and a longitudinal design are recommended to establish causality.

## Conclusion

Increased risk to obstructive sleep apnea among young adults is associated with acute stress, and the relationship is mediated by sleep quality.

## Data Availability

The data is available upon reasonable request.

## Acknowledgments

We would like to acknowledge the Indian Council of Medical Research. This study was conducted under the Short-Term Studentship Award (STS-2019) conferred by the Indian Council of Medical Research.

## Disclosure

The authors report no conflicts of interest in this work.

The abstract of this paper was presented at the 15^th^ World Sleep Congress 2019, September 21-25, at Vancouver, Canada as a poster presentation with interim findings. The poster’s abstract was published in “Poster Abstracts” in the journal ‘Sleep medicine’ and is available at https://www.sciencedirect.com/journal/sleep-medicine/vol/64/suppl/S1.

